# Epidemiology of first-and second-line anti-tuberculosis drug resistance in new pulmonary tuberculosis cases in Addis Ababa metropolitan area, Ethiopia

**DOI:** 10.1101/2023.04.20.23288854

**Authors:** Gizachew Taddesse Akalu, Belay Tessema, Waganeh Sinshaw, Misikir Amare, Getu Diriba, Melak Getu, Betselot Zerihun, Beyene Petros

## Abstract

**Background:** Conventional wisdom wrongly holds that the microbiological of *M. tuberculosis* complex in clinical specimens via culture and phenotypic drug susceptibility testing allows people to be correctly diagnosed and ensures an effective treatment regimen to be selected. This study was aimed to characterize first-and second-line anti-tuberculosis drug resistance profiles among new pulmonary tuberculosis cases in Addis Ababa metropolitan area, Ethiopia.

**Methods:** A prospective cross-sectional study was conducted between October 2019 and June 2021 among bacteriologically confirmed new presumptive pulmonary tuberculosis cases. GeneXpert MTB/RIF Assay was utilized for initial testing and early detection of rifampicin resistance. Mycobacterial culture and drug susceptibility testing were performed against FOUR first-line and ELEVEN second-line anti-TB drugs using BD BACTEC™ MGIT™ 960 automated liquid culture system.

**Results:** A total of 156 *M. tuberculosis* complex isolates were successfully recovered using BD BACTEC™ MGIT™ 960 automated liquid culture system and were subjected to drug susceptibility testing. Males account for 53.8 % (84/156). The median age of the study participants was 30.0 years. Of all the study participants, 58.3 % (91/156) were married, and 76.9% (120/156) were urban residents. Overall, we identified 14.1% (22/156) resistance to at least one anti-TB drug and 85.9% (134/156) pan-susceptible *M. tuberculosis* strains. Further, 7.1% (11/156) of isolates were monoresistant, 5.8% (9/156) of isolates were MDR-TB strains, and 3.8% (6/156) of isolates were resistant to all first-line anti-TB drug regimens. Interestingly, all isolates were susceptible to all recently recommended second-line anti-TB drugs, and none of these isolates were found to be pre-XDR or XDR-TB. The rate of RR-TB detected was 10.9% (17/156) and 5.8% (9/156) using GeneXpert MTB/RIF Assay and BD BACTEC™ MGIT™ 960 SIRE liquid culture system, respectively. The sensitivity, specificity, PPV, NPV, accuracy, and Kappa value were 100%, 94.6%, 52.9%, 100%, 94.9%, and 0.667, respectively.

**Conclusion:** The rate of MDR-TB in new pulmonary TB cases remained high at fivefold the national and nearly twofold the global estimated rate. The rate of monoresistance against anti-TB drugs was also high. The absence of resistance against recommended second-line anti-TB drugs was quite encouraging. However, the high rate of resistance against Ethionamide would mean that its inclusion in the regimens may not have therapeutic benefit in this geographic area. Furthermore, the low specificity of GeneXpert MTB/RIF Assay might introduce a significant rate of (47.1%; 8/17) false rifampicin resistance leading the patient to erroneous MDR-TB category and placing on an unnecessary second-line anti-TB-treatment regimen. Enhanced efforts are required to progressively validate and harmonize rapid molecular diagnostics against reference methods to address the diagnosis challenges and improve patient outcomes.

## Introduction

Tuberculosis (TB), caused by mycobacterium tuberculosis (*M. tuberculosis*) is the oldest disease known to affect humans with myriad clinical manifestations of diseases and fatalities in the world [1-4]. *Mycobacterium. tuberculosis* belongs to closely related mycobacterial species including *M. bovis, M. africanum, M. microti, M. caprae, M. pinnipedii, M. canetti*, and *M. mungi*, together comprising *M. tuberculosis* complex [5 - 7]. The origin and global spread of human TB had a contemporary notion that the genus Mycobacterium originated in animals and was transferred to humans during the neolithic transition 150 million years ago [1, 8, 9]. Recent evolutionary data suggested that *M. tuberculosis* emerged as a human pathogen in Africa and survived with modern human migrations approximately 70 000 years ago [4,10,11]. However, it was in 1882, when Robert Koch was able to isolate tubercle bacillus that determine a milestone in the fight against TB [12-14].

According to the WHO global tuberculosis report of 2022, an estimated 10.6 million persons became ill from TB worldwide in 2021, of which 5.3 million people were diagnosed with pulmonary TB. Similarly, an estimated 1.6 million deaths and an estimated 4 million people with TB missed undiagnosed in 2021[15]. This reversal of progress from the previous estimations is indicative of challenges in closing the gaps to end TB. The emergence of drug resistance *M. tuberculosis* strains against commonly used and less toxic standard anti-TB drugs continue to be a clear threat to the realization of TB elimination [16,17]. Globally, 3.6% of new TB cases and 18% of previously treated cases had MDR or RR-TB. Conversely, about 2 billion people are estimated to generate a latent TB infection, one of the most remarkable features of *M. tuberculosis*, and are thus at risk of developing active TB disease in their lifetime [4,15,18]. Ethiopia still remained among the 30 TB and TB/HIV high-burden countries, while transitioning out of the list of MDR/RR-TB for 2021 – 2025. According to the WHO global tuberculosis report in 2022, Ethiopia constitutes an annual TB incidence rate of 143 per 100,000 population. Similarly, an estimated proportion of 1.1% and 12% MDR/RR-TB in new and in previously treated TB cases were reported respectively [15].

Globally, a remarkable campaign to control tuberculosis was started by WHO in 1948, and in 1974 the WHO Expert Committee on Tuberculosis issued a policy guideline to control TB by discouraging both radiographic screening and tuberculin skin testing while promoting sputum microscopy and BCG vaccination [14]. Following years of increased incidences, the WHO declared TB a global emergency in 1993 and in 1995 devised a directly observed treatment (DOTs) as a control strategy with the aspiration to save lives and dream of TB elimination [13,19]. Efforts to control TB in Ethiopia began with the sequential entrenchment of TB centers in the early 1960s, the National Tuberculosis Control Program central office in 1976, and the National Tuberculosis and Leprosy Control Program in 1994. The country adopted the DOTs strategy in 1997 and endorsed the ‘Stop TB’ and ‘End TB’ aspirational strategies of the UN MDGs and SDGs [20, 21]. However, despite concerted efforts during the past decades, the rate of decline in tuberculosis incidence, and mortality is lower than what would be required to reach the local as well as the global target of pre-elimination by 2035 and elimination by 2050.

The microbiological detection of TB is critical as it allows people to be correctly diagnosed, is necessary for drug resistance detection, and ensures that the most effective treatment regimen can be selected as early as possible. Tuberculosis is mostly diagnosed using smear microscopy, rapid molecular methods, culture, and phenotypic and genotypic DST methods. However, the gold standard method for the bacteriological confirmation of TB and detecting resistance to anti-TB drugs is culture using commercially available liquid media [22, 23].

The standard treatment regimens for susceptible new TB patients are 2RHZE:4RH divided into intensive and continuation phases respectively. While, the updated standard shorter MDR-TB and longer MDR-TB regimen to treat MDR/RR-TB are recently adopted by the MoH as follows: 4–6 BDQ -LFX-CFZ-Z-E-HH-ETO:5 LFX-CFZ-Z-E; and 18 BDQ: LFX/MFX-LZD-CFZ-CS respectively [15, 18, 21,24].

Nonetheless, efficient monitoring response to ant-TB drugs is quite critical to ensure that patients are responding to treatment at an acceptable success rate, to guide policymakers, to effectively limit transmission of resistant strains in the community, to improve the quality of life of every TB patient and to recognize *M. tuberculosis* drug resistance pattern in a specific geographic area. However, monitoring responses to ant-TB drugs using molecular assay is not suitable, and culture and phenotypic DST is not routinely utilized in Ethiopia. Furthermore, while it is quite important to assign patients to appropriate treatment regimens, evidence of the agreement between phenotypic and WHO-recommended rapid molecular drug resistance detection is elusive. The aim of this study was to describe the epidemiology of first-and second-line anti-TB drug resistance situations in new presumptive pulmonary tuberculosis cases in Addis Ababa metropolitan area, Ethiopia.

## Materials and Methods

### Study design and setting

A prospective cross-sectional study was conducted between October 2019 and June 2021 among clinically eligible new presumptive pulmonary TB cases recruited at Saint Peter TB Specialized Hospital, a national reference TB treatment initiation center in Addis Ababa metropolitan city, and from adjacent towns of Sululta and Sendafa Health Centers. All clinically suspected cases were bacteriologically confirmed positive MTB patients using GeneXpert MTB/RIF assay triage testing and this was performed based on the national guideline and as recommended by WHO for screening and diagnosis of TB. Mycobacterial culture and phenotypic drug susceptibility testing were performed using BD BACTEC™ MGIT™ 960 automated mycobacterial detection liquid culture system, Becton Dickinson, Sparks, MD, USA, at the National TB Reference Laboratory of Ethiopia Public Health Institute, Addis Ababa, Ethiopia. Prior to sputum specimen collection, sociodemographic and clinicoepidemiological data were captured using a structured and validated tool prepared for this study. Data were delinked from the source file, avoided study subject identifiers, and analyzed in aggregates to ensure confidentiality and anonymity.

### Study participants

The study population was all clinically suspected new pulmonary TB cases, bacteriologically confirmed with GeneXpert MTB/RIF assay on triage testing and further confirmed with BD BACTEC™ MGIT™ automated mycobacterial detection solution for mycobacterial liquid culture and susceptibility testing. The drug resistance patterns were the outcome variables of this study, whereas the sex, age, marital status, occupation, residence, religion, previous history related to TB symptoms, and behavior-related risk factors including perceived HIV status, intravenous drug use, tobacco smoking, alcohol use, and khat use were the independent variables which could determine factors associated with DST pattern and were investigated in this study.

### Inclusion and exclusion criteria

The inclusion criteria were new clinically suspected presumptive pulmonary TB cases whose suspicion were bacteriologically confirmed positive for MTB infection with GeneXpert MTB/RIF assay were considered for enrollment in this study. The exclusion criteria were being on treatment for TB within the last three months before the commencement of this study, treated for TB or has taken anti-TB medicines for more than one month, extrapulmonary TB suspects who were unable to provide sputum, and those who refused to participate were avoided from enrollment in this study. Furthermore, the WHO case definition was applied, in which a new presumptive pulmonary TB case was defined as a newly identified episode of TB case who has never been treated for TB or has taken anti-TB medicines for less than a month.

### Sample collection, transport, and storage

All clinically eligible newly recruited presumptive pulmonary TB cases who voluntarily demonstrated written informed consent were demanded to provide two sputum samples of 3-5ml each in a sterile falcon tube of 50 ml capacity in front of a laboratory technologist. The first sputum specimens were collected upon enrollment and were analyzed using GeneXpert MTB/RIF assay without any delay, while the second sputum specimens were collected in the morning the following day for isolation of TB bacilli and DST. Both sputum specimens were expectorated and without mechanical maneuver. Each patient’s second sputum specimen was stored at 2 - 8 ^0^C at St. Peter TB Specialized Hospital for a maximum of 24 hours after collection. Using appropriate packaging and a reverse cold chain system, all specimens were transported to the National Tuberculosis Reference Laboratory at the Ethiopian Public Health Institute for isolation of tubercle bacilli and drug susceptibility testing using BD BACTEC™ MGIT™ 960 automated mycobacterial detection system.

### Laboratory procedures

#### GeneXpert MTB/RIF Assay triage testing

The GeneXpert MTB/RIF assay was performed according to the manufacturer’s instruction, Cepheid, Sunnyvale, CA, USA. It is an automated in vitro diagnostic test using nested real-time PCR for the qualitative and semiquantitative detection of *M. tuberculosis* complex and simultaneous RR TB. Generally, results from GeneXpert MTB/RIF assay were obtained and assessed with four result types namely: MTB Detected, MTB Detected RR-TB Detected, MTB Not Detected, and Invalid/Error/No result.

#### Digestion, decontamination, and concentration of Sputum for culture and DST

Sputum specimens were digested and decontaminated by the standard NALC-NaOH method as recommended by the WHO and GLI mycobacteriology laboratory manual using BD BBL™ MycoPrep™ Kit [25]. Sodium citrate and phosphate buffer were used to exert the stability of NALC and neutralize the NaOH homogenate respectively. The supernatant was discarded and the pellet was resuspended with sterile phosphate-buffered saline (PBS). The mixture was used for inoculation of the BD BACTEC™ MGIT™ 960 tubes. Prior to specimen inoculation, 0.5ml of OADC (Oleic acid, albumin, dextrose, and catalase) growth enrichment and 0.1ml of PANTA (polymyxin B, amphotericin B, nalidixic acid, trimethoprim, and azlocillin) antibiotic mixture was added to the Mycobacteria Growth Indicator Tube that contains 4 mL of modified Middlebrook 7H9 Broth base. Finally, the MGIT tubes were inoculated with 0.5 ml of the concentrated specimen suspension, tightly recapped, mixed well, and incubated at 37 °C in the BD BACTEC™ MGIT™ 960 instrument system in which tubes were automatically monitored each hour for fluorescence development for 42 days or until a positive signal developed.

#### Isolation and identification of mycobacteria species

Sputum specimens were processed and cultured according to the national TB reference laboratory, Ethiopian Public Health Institute and WHO recommended protocol using BD BACTEC™ MGIT™ 960 automated liquid culture system, Becton Dickinson, Sparks, MD, USA. A brain heart infusion agar plate was used to check for contamination, and a media smear for AFB was performed on all positive growths to confirm tubercle bacilli microscopically. To discriminate Mycobacterium tuberculosis complex from Mycobacteria other than tubercle bacilli, isolates were identified by TB Antigen MPT64 Rapid Test, SD Bioline, Republic of Korea. All mycobacteria isolates were stored at -80^0^C until DST was performed.

#### First- and second-line drug susceptibility testing

Phenotypic DST for first-line anti-TB medicines was performed using the BD BACTEC™ MGIT™ 960 SIRE Kits for the antimycobacterial susceptibility testing of *Mycobacterium tuberculosis* from culture. The critical concentrations of drugs used were as follows: 2.0μg/ml for STM; 0.1μg/ml for INH; 1.0μg/ml for RIF and 5.0μg/ml for EMB as recommended by WHO and previously described by the Global Laboratory Initiative [26, 27]. The DST was not performed for PZA. Whilst, phenotypic DST for second-line anti-TB medicines include fluoroquinolones (Levofloxacin, Moxifloxacin, and Ofloxacin), aminoglycoside injectables (Amikacin, Capreomycin, and Kanamycin), and other core second-line agents (Ethionamide, Bedaquiline, Clofazimine, Delamanid, and Linezolid). The critical concentrations of drugs used were: 1.0 μg/ml for Amikacin, 2.5 μg/ml for Capreomycin, 2.5 μg/ml for Kanamycin, 1.0 μg/ml for Levofloxacin, 0.5 μg/ml for Moxifloxacin, 2.0 μg/ml for Ofloxacin and 5.0 μg/ml for Ethionamide. Furthermore, the critical concentration for WHO-recommended medicines and recently optimized were included as follows: 1.0 μg/ml for Bedaquiline, 1.0 μg/ml for Clofazimine, 0.06 μg/ml for Delamanid and 1.0 μg/ml for Linezolid as recommended by WHO and previously described by the Global Laboratory Initiative [26, 27]. The MGIT 960 system monitors these growth patterns and can automatically interpret results as susceptible or resistant. An isolate is defined as resistant if 1% or more of the test population grows in the presence of the critical concentration of the drug. *Mycobacterium tuberculosis* strain H37Rv was used as a sensitive control for the susceptibility testing.

### Data analysis

Data were entered in Microsoft Excel 2022, checked for inconsistencies, and cleaned promptly. The collected data were exported to IBM SPSS Statistics for Windows, Version 26.0. (Armonk, NY: IBM Corp. USA) for analysis. Descriptive parameters were used to explain the study subject’s clinical profiles and drug susceptibility patterns. Mean and standard deviation was used to depict continuous variables. Statistical significance was determined to estimate the precision at a 95% confidence interval. The results were presented in tables and figures as appropriate.

### Quality assurance

To ensure the quality of the samples, good laboratory practices, and to maintain a high standard of accuracy of results, standard operating procedures were strictly followed during sputum specimen collection, transportation, processing, and laboratory analysis. The internal quality control of the GeneXpert MTB/RIF assay system was validated using non-RR and known RR *M. tuberculosis* H37Rv strains stored at -20°C. The GeneXpert MTB/RIF assay system has inbuilt internal quality monitoring systems to control adequate processing of the target bacteria, monitor the presence of inhibitors in the PCR reaction, to verifies reagent rehydration, PCR tube filling in the cartridge, probe integrity, and dye stability. The sterility of the culture media was checked by incubating the whole media at 37°C for 48 hours and the performance was checked by known drug-susceptible *M. tuberculosis* H37Rv reference strain ATCC 27294. The sterility of sample processing reagents was checked by inoculating all reagents on BHI. Positive and negative controls were included in each batch of culture and DST. Sterile molecular grade water and reagent control were used as a negative control, while H37Rv ATCC25177 was used as a positive control. All laboratory results were recorded on a separate logbook prepared for this purpose during the study period. Moreover, all specimens suspected of containing *M. tuberculosis* were handled with appropriate precaution at all times and opened only within an appropriate biosafety cabinet.

## Results

### sociodemographic and clinical characteristics

*M. tuberculosis* was isolated from 216 new presumptive pulmonary TB cases with MTB-positive GeneXpert MTB/RIF Assay results diagnosed between October 2019 and July 2020. DST was performed till June 2021. We excluded 60 patients from the analysis based on the fidelity of liquid culture, mainly due to viability and contamination. Overall, 15.7% (34/216) and 12.0% (26/216) of specimens were found to be not viable and contaminated respectively (Fig 1).

**Figure 1:**
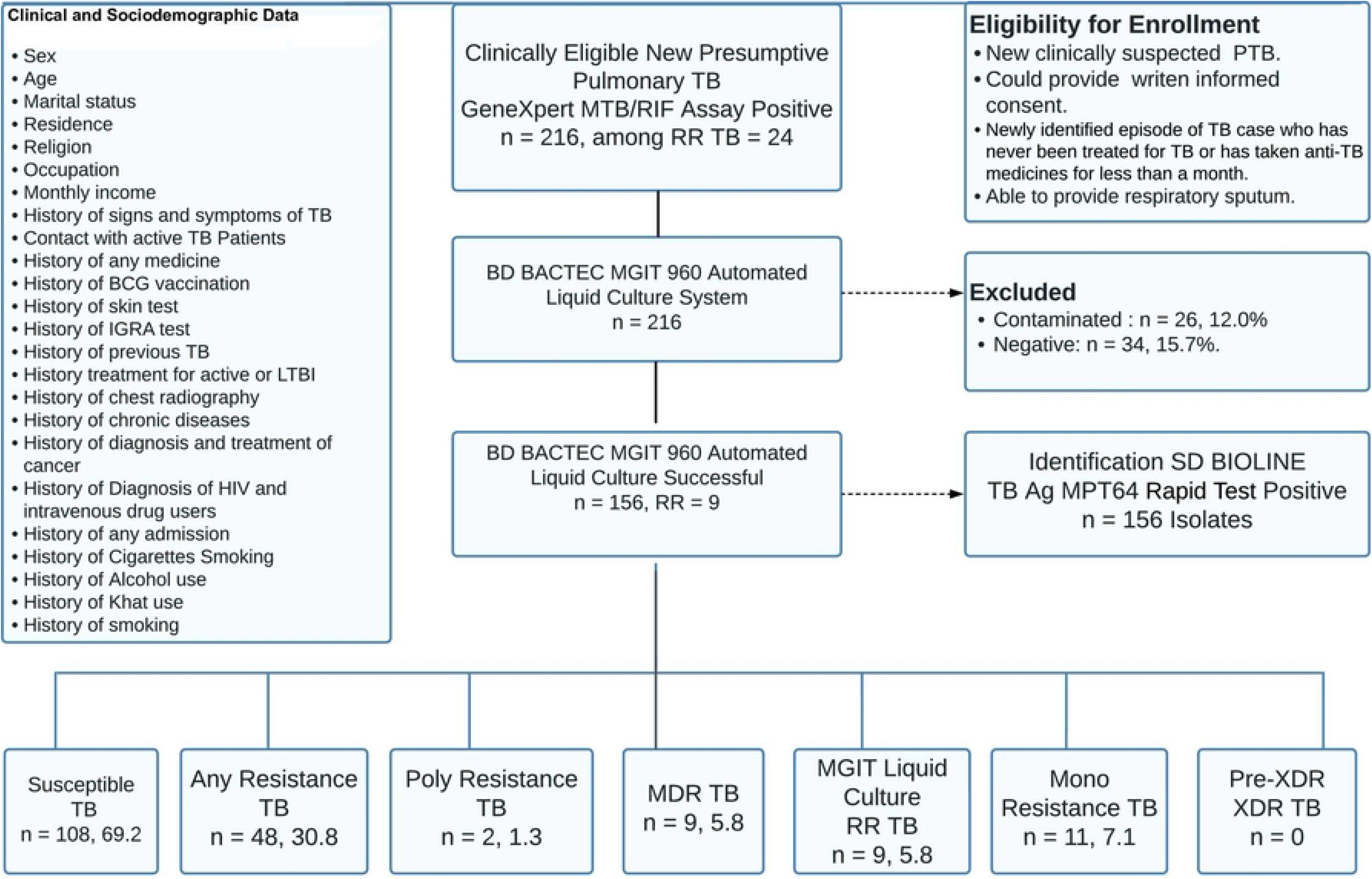
Flowchart showing clinical and experiement workflow.

A total of 156 *Mycobacterium tuberculosis* complex isolates were successfully recovered using a gold standard BD BACTEC™ MGIT™ 960 automated liquid culture system and were subjected to drug susceptibility testing. Among the study participants, males account for 53.8 % (84/156). The median age of the study participants was 30.0 (range 6 – 75) years with a mean and standard deviation of 33.56 and 12.65 years, respectively. Of all the study participants, 49.4 % (77/156) were in the age group below 30.0 years, 58.3 % (91/156) were married, and 76.9% (120/156) were urban residents (Table 1).

**Table 1.** Characteristics of the study participants in Addis Ababa metropolitan area, Ethiopia, (n = 156)

| Variables Measured | Classification | Frequency Distribution |  |
| --- | --- | --- | --- |
|  |  | Number [n] | Percent [%] |
| Patients by health facility | SPH<sup>a</sup> | 100 | 64.1 |
|  | SulHC<sup>b</sup> | 25 | 16.0 |
|  | SenHC<sup>c</sup> | 31 | 19.9 |
| Sex | Male | 84 | 53.8 |
|  | Female | 72 | 46.2 |
| Age group | < = 29 Years | 77 | 49.4 |
|  | 30 – 50 Years | 59 | 37.8 |
|  | > =51 Years | 20 | 12.8 |
| Marital status | Single | 60 | 38.5 |
|  | Married | 91 | 58.3 |
|  | Divorced | 5 | 3.2 |
| Residence | Urban | 120 | 76.9 |
|  | Rural | 36 | 23.1 |
| Religion | Christian | 130 | 83.3 |
|  | Muslim | 26 | 16.7 |
| Occupation | House Wife | 23 | 14.8 |
|  | Daily Laborer | 57 | 36.5 |
|  | Employed | 52 | 33.3 |
|  | Unemployed | 24 | 15.4 |
| Monthly income in ETB | < = 500 | 74 | 47.4 |
|  | 501 – 1,999 | 33 | 21.2 |
|  | 2,000 -10,000 | 49 | 31.4 |
| Previous history of TB symptom | Yes | 154 | 98.7 |
|  | No | 2 | 1.3 |
| History of contact with people with active TB | Yes | 48 | 30.8 |
|  | No | 108 | 69.2 |
| History of any medicine | Yes | 38 | 24.4 |
|  | No | 118 | 75.6 |
| History of BCG vaccination | Yes | 25 | 16.0 |
|  | No | 131 | 84.0 |
| History of TB skin test | Yes | 16 | 10.3 |
|  | No | 140 | 89.7 |
| History of TB IGRA test | Yes | 20 | 12.8 |
|  | No | 136 | 87.2 |
| History of previous PTB+ | Yes | 27 | 17.3 |
|  | No | 129 | 82.7 |
| Treatment history for active or LTBI | Yes | 34 | 21.8 |
|  | No | 122 | 78.2 |
| History of CXR for pulmonary TB | Yes | 92 | 59.0 |
|  | No | 64 | 41.0 |
| History of chronic diseases | Yes | 34 | 21.8 |
|  | No | 122 | 78.2 |
| Previous Rx and Dx history for CA | Yes | 6 | 3.8 |
|  | No | 150 | 96.2 |
| History of HIV and IDUs | Yes | 37 | 23.7 |
|  | No | 119 | 76.3 |
| History of hospital admission | Yes | 24 | 15.4 |
|  | No | 132 | 84.6 |
| History of alcohol consumption | Yes | 39 | 25.0 |
|  | No | 117 | 75.0 |
| History of khat consumption | Yes | 30 | 19.2 |
|  | No | 126 | 80.8 |
| History of smoking | Yes | 21 | 13.5 |
|  | No | 135 | 86.5 |

**ETB:** Ethiopian Birr; **TB:** Tuberculosis; **\*SPH:** Saint Peter Hospital; **<sup>b</sup>SulHC:** Sululta Health Center; **<sup>c</sup>SenHC:** Sendafa Health Center; **BCG:** Bacille Calmette-Guerin; **IGRA:** Interferon-Gamma Release Assay; **PTB+:** Pulmonary TB Positive; **CXR:** Chest X-Ray; **Rx:** Treatment; **Dx:** Diagnosis; **CA:** Cancer; **HIV:** Human Immunodeficiency Virus; **IDUs:** Intravenous Drug Users.

### Phenotypic DST and characteristics of isolates against first-line anti-TB drugs

Overall, from the 156 *M. tuberculosis* complex isolates, we identified 14.1% (22/156) resistance to at least one anti-TB drug, while 85.9% (134/156) of *M. tuberculosis* strains were identified as pan-susceptible. Further, 7.1% (11/156) of isolates were monoresistant *M. tuberculosis* strains, 5.8% (9/156) of isolates were MDR-TB strains, and 3.8% (6/156) of isolates were identified as resistant to all first-line anti-TB drugs tested. Resistance to any first-line anti-TB drugs was 9.0% (14/156) for STM, 11.5% (18/156) for INH, 5.8% (9/156) for RIF, and 4.5% (7/156) for EMB. From the 7.1% (11/156) of monoresistant isolates against first-line anti-TB drugs, 5.1% (8/156) were INH monoresistant and 1.9 % (3/156) were STM monoresistant. A resistance profile to PZA was not available in this study. Interestingly, monoresistance to RIF and EMB was not demonstrated in all isolates (Table 2).

**Table 2.** Phenotypic drug susceptibility testing profile to first- and second-line anti-TB drugs (n = 156)

| Drug Profile and Regimen Category | Frequency Distribution |  | Type of Drug Resistance TB |
| --- | --- | --- | --- |
|  | Susceptible n [%] | Resistant n [%] |  |
| First Line Drug Profile |  |  |  |
| All isolates with Resistance Profile | 134 [85.9] | 22[14.1] | Any Resistant TB |
| STM | 142 [91.0] | 14[9.0] |  |
| INH | 138 [88.5] | 18[11.5] |  |
| RIF | 147 [94.2] | 9[5.8] |  |
| EMB | 149 [95.5] | 7[4.5] |  |
| STM+INH | 155 [99.4] | 1 [0.6] | Poly Resistant TB |
| STM+INH+EMB | 155 [99.4] | 1 [0.6] |  |
| Total |  | 2[1.3] |  |
| INH | 148 [94.9] | 8 [5.1] | Mono Resistant TB |
| STM | 153 [98.1] | 3 [1.9] |  |
| Total |  | 11[7.1] |  |
| INH+RIF | 155 [99.4] | 1 [0.6] | Multidrug Resistant TB |
| INH+RIF+STM | 154 [98.7] | 2 [1.3] |  |
| INH+RIF+STM+EMB | 150 [96.2] | 6 [3.8] |  |
| Total |  | 9[5.8] |  |
| Second-Line Drug Profile |  |  |  |
| MDR+AMK | 22 [100] | 0 | Pre-XDR |
| MDR+CAP | 22 [100] | 0 |  |
| MDR+KAN | 22 [100] | 0 |  |
| MDR+LFX | 22 [100] | 0 |  |
| MDR+MFX | 22 [100] | 0 |  |
| MDR+OFX | 22 [100] | 0 |  |
| MDR+BDQ | 22 [100] | 0 |  |
| MDR+CZF | 22 [100] | 0 |  |
| MDR+DLM | 22 [100] | 0 |  |
| MDR+LZD | 22 [100] | 0 |  |
| MDR+ETO<sup>a</sup> | 4[18.2] | 18 [81.8] |  |
| MDR+LFX+MFX+OFX | 21 [100] | 0 | XDR |
| MDR+LFX+MFX+BDQ+LZD | 21 [100] | 0 |  |

**MDR:** Multidrug-resistant; **STM:** Streptomycin; **INH:** Isoniazid; **RIF:** Rifampicin; **EMB:** Ethambutol; **AMK:** Amikacin; **CAP:** Capreomycin; **KAN:** Kanamycin; **LFX:** Levofloxacin; **MFX:** Moxifloxacin; **OFX:** Ofloxacin; **BDQ:** Bedaquiline; **CFZ:** Clofazimine; **DLM:** Delamanid; **LZD:** Linezolid; **ETO:** Ethionamide.
<sup>a</sup>ETO: This is not among the recommended drugs, but could be conditionally prescribed.

### Phenotypic DST and characteristics of isolates against second-line anti-TB drugs

Phenotypic drug susceptibility testing against second-line anti-TB agents that include fluoroquinolones: Levofloxacin, Moxifloxacin, and Ofloxacin; aminoglycosides: Amikacin, Capreomycin, and Kanamycin; and other core second-line agents: Ethionamide, Bedaquiline, Clofazimine, Delamanid, and Linezolid was performed using BD BACTEC™ MGIT™ 960 instrument system. Among all drug-resistant strains (any), 81.8% (18/22) of isolates were resistant to ETO. However, all *M. tuberculosis* isolates that demonstrated any resistance to at least one anti-TB first-line drug tested were found susceptible to WHO-recommended and nationally endorsed second-line anti-TB drugs (Table 2).

### RR-TB detected in GeneXpert Vs. BD BACTEC MGIT 960 liquid culture system

In this study, the rate of RR-TB detected was 10.9% (17/156) and 5.8% (9/156) using GeneXpert MTB/RIF Assay and BD BACTEC™ MGIT™ 960 SIRE liquid culture system, respectively. Further, rifampicin resistance detected using GeneXpert MTB/RIF Assay compared to BD BACTEC™ MGIT™ 960 SIRE liquid culture system for automated DST showed a sensitivity of 100%, specificity of 94.6%, positive predictive value (PPV) of 52.9%, and negative predictive value (NPV) of 100% (Table 3).

**Table 3.** RR-TB detection rate among GeneXpert MTB/RIF Assay and BACTEC™ MGIT™ 960 automated liquid culture system.

| Methods Used |  | RIF Resistance Detected in BD BACTEC 960 Liquid Culture System |  |  |  |  |  |  |  |  |  |  |
| --- | --- | --- | --- | --- | --- | --- | --- | --- | --- | --- | --- | --- |
|  |  | R | S | Total | Sensitivity (%) | Specificity (%) | PPV (%) | NPV (%) | Accuracy (%) | 95% CI | Kappa Value | P. value, 95% CI |
| GeneXpert MTB/RIF Assay | R | 9 | 8 | 17 | 100 | 94.6 | 52.9 | 100 | 94.9 | 90.2 - 97.8 | 0.667 | <0.001 [0.284-0.779] |
|  | S | 0 | 139 | 139 |  |  |  |  |  |  |  |  |
| Total |  | 9 | 147 | 156 |  |  |  |  |  |  |  |  |

**R:** Resistant; **S:** Sensitive; **PPV:** Positive Predictive Value; **NPV:** Negative Predictive Value; **CI:** Confidence Interval.

## Discussion

In this study, we identified a rate of 14.1% (22/156) resistance of *M. tuberculosis* strains to at least one first-line anti-TB drug, while 85.9% (134/156) were found to be pan-susceptible *M. tuberculosis* strains in new presumptive pulmonary TB cases. Similar to this finding, a comparable rate of resistance of *M. tuberculosis* strains to at least one first-line anti-TB drug among new cases was reported by Tessema B et al., (15.8%, 2012) [28], Adane K et al., (15.58%, 2015) [29], Lobie TA et al., (16.1%, 2020) [30], Worku G et al., (11.6%, 2022) [31] in Ethiopia; Doss M et al., (13.4%, 1999) [32] in Côte d’Ivoire; Antunes ML et al., (17.8%, 2000) [33] in Portugal; Glasauer S et al., (12.7%, 2019) [34] in Germany and Ullah I et al., (11.5%, 2016) [35] in Pakistan. Compared to our finding, a higher rate of resistance of *M. tuberculosis* strains to at least one first-line anti-TB drug among new cases was reported by Yimer SA et al., (30.1%, 2012) [36], Bedewi Z et al., (22.2%, 2017) [37], Seyoum B et al., (23.0%, 2015) [38], Dagne B et al., (26.8%, 2021) [39], Yigzaw WB et al., (34%, 2021) [40] in Ethiopia; Mende N et al., (23.5%, 2023) [41] in Zambia; Thapa G et al., (31.1%, 2016) [42] in Nepal; Li Y et al., (34.1%, 2016) [43] in China; Ndung’u PW et al., (30.0%, 2012) [44], Kerubo G et al., (30.0%, 2016) [45] and Yonge SA et al., (48.6%, 2017) [46] in Kenya; and Aung WW et al., (27.7%, 2015) [47] in Myanmar. However, a lower rate of resistance of *M. tuberculosis* strains in new presumptive pulmonary TB cases was reported in the previous studies by Biadglegne F et al., (6.7%, 2014) [48] in Ethiopia.

The most likely justification for these differences in the rate of drug resistance reported by the previous studies in the same as well as different geographic regions might be due to the difference in the study period, study design, study subject, sample size, health system efficiency to TB control activities, patient’s immunological and economic status and virulence factors of the Mycobacterial strain circulating in the community.

The rate of MDR-TB identified in this study was 5.8%, (9/156) among new presumptive TB cases and this was congruent to the rate of MDR-TB reported in the previous studies by Tessema B et al., (3.7%, 2012) [28], Yigzaw WB et al., (5.0%, 2021) [40] in Ethiopia; Yonge SA et al., (4.8%, 2017) [46] in Kenya and Dosso M et al., (5.3%, 1999) [32] in Côte d’Ivoire. However, compared to our study findings, a higher rate of MDR-TB was reported in the previous studies by Welekidan LN et al., (11.6%, 2020) [49], and Dagne B et al., (61.9%, 2021) [39] in Ethiopia; Ullah I et al., (9.3%, 2016) [35] in Pakistan; Monde N et al., (9.8%, 2023) [41] in Zambia, Thapa G et al., (15.6%, 2016) [42] in Nepal, Li Y et al., (13.2%, 2016) [43], and Zhou L et al., (18.8%, 2022) [50] in China and Aung WW et al., (18.3%,2015) [] in Myanmar. On the other hand, a lower rate of MDR-TB was reported by Yimer SA et al., (1.0%, 2012) [36], Seyoum B et al., (1.1%, 2015) [38], Bedewi Z et al., (1.5%, 2017) [37], Adane K et al., (1.29%, 2015) [29], Mekonnen F et al., (2.3%, 2015) [51], and Lobie TA et al., (2.3%, 2020) [30] in Ethiopia; Glasauer S et al., (3.3%, 2019) [34] in Germany. Notwithstanding this observation, the WHO estimate of 1.1% of MDR-TB in new TB cases revealed in the 2022 global tuberculosis report is much lower than the findings demonstrated in this study [15].

The most plausible justification for the higher rate of MDR-TB among new presumptive pulmonary TB cases in our study than in the previous studies may contemplate the emergence and transmission of multi-drug resistant TB in Ethiopia remain a challenge and shows that the battle against TB is far from over within the anticipated local as well as global initiatives. Furthermore, in addition to the biological factors leading to the high prevalence of MDR-TB, such as the fitness of DR-MTB strains, mutation ability, and virulence variation, the rate of poor treatment adherence and inefficient health service system have a significant influence on the development of MDR-TB. Nevertheless, the DOTS strategy has been proven to be one of the most effective strategies to manage TB and limit the emergence and transmission of MDR-TB in the community. However, despite the tremendous efforts and encouraging progresses in widening DOTs coverage in Ethiopia in public as well as private TB treatment initiation centers, evidence showed active TB case surveillance, as well as rapid detection of drug resistance, remained low to end TB by 2035 and eliminate TB by 2050 partly because culture and universal DST are not widely accessible and routinely done in Ethiopia.

In this study, 3.8% (6/156) of isolates were identified as resistant to all first-line anti-TB drugs tested, including STM, INH, RIF, and EMB. This proportion of resistance was higher than the reported proportion in the previous study done by Yigzaw WB et al., (2.38%, 2021) [40] in Ethiopia. In addition, 7.1% (11/156) of isolates were mono-resistant *M. tuberculosis* strains, and the highest mono-drug resistance isolates to first-line anti-TB drugs were from INH (5.1%, 8/156) and STM (1.9%, 3/156). A congruent monoresistance rate was reported in the previous study by Zürcher K et al., (7.0%, 2019) [52] in seven high TB-burden countries. This rate of INH and STM monoresistance respectively was lower than the reported rate in the previous studies by Seyoum B et al., (9.5%, 7.0%, 2015) [38] and Bedewi Z et al., (6.3% and 7.5%, 2017) [37] in Ethiopia. However, compared to our study findings, variably a higher rate of INH and a lower rate of STM monoresistance were reported in the previous study by Monde N et al., (9.8% and 0.8%, 2023) [41], respectively, in Zambia.

This study further demonstrated that the rate of resistance to any of the first-line anti-TB drugs to be 9.0% (14/156), 11.5% (18/156), 5.8% (9/156), and 4.5% (7/156) for STM, INH, RIF, and ETM respectively. A comparable rate of any resistance to first-line anti-TB drugs was reported in the previous studies by Seyoum B et al in 2015 [38], and Adane K et al., in 2015 [29] in Ethiopia, and an incomparably higher rate of drug resistance (any) related to INH and RIF was reported in the previous study by Thapa d et al., (23.0% and 17.8%, 2016) [42], respectively, in Nepal. Interestingly, monoresistance to RIF and EMB was not demonstrated in all isolates in this study. The absence of monoresistance against RIF and EMB was reported in the previous studies by Agonafir M et al., (2010) [53], Tessema B et al., (2012) [28], and Worku G et al., (2022) [31] in Ethiopia. A higher rate of sensitivity to RIF alone may be a good indicator of the success of the DOTs program in the study area in Ethiopia. However, the higher rate of INH monoresistance observed in new presumptive TB cases in this study is critical as INH is the main monotherapy drug used to treat LTBIs and the main chemoprophylaxis drug used in immunosuppressed TB patients in Ethiopia.

The findings of this study demonstrated that all MDR-TB isolates were susceptible to all recently recommended second-line anti-TB drugs tested and none of these isolates were found to be Pre-XDR or XDR-TB. Compared to this finding, similar results were reported in previous studies conducted by Tessema B et al., (2012) [28], and Worku G et al., (2022) [31] in Ethiopia. However, a significant proportion (81.8%) of isolates exhibited resistance to Ethionamide in this study. Although higher rate resistance against second-line anti-TB drugs was reported in the previous studies by Agonafir M et al., (73.9%, 2010) [53], and Dagne B et al., (5.6%, 2021) [40] in Ethiopia, and Monde N et al., (35.5%, 2023) [41] in Zambia, the absence of resistance to appropriate second-line anti-TB drugs in this study may reflect that the existing treatment of MDR-TB cases in the study area was encouragingly effective. This observation is critical because of the positive implication that initiation of second-line drugs without initial DST might not be beneficial to the patient and requires reemphasizing the need to perform drug susceptibility testing prior to granting anti-TB treatment. Nevertheless, previous studies reported that diagnosed, untreated, and inappropriately managed MDR-TB contributes to a continuously sustained high prevalence of drug-resistant *M. tuberculosis* strains circulating in the community [54].

However, compared to this study finding, a high rate of resistance against Ethionamide was reported in the previous studies by Ongaya VA et al., (56.3%, 2012) [55] in Kenya, Rueda J et al., (95.0%, 2017) [56] in Cambodia and Islam MM et al., (90.8%, 2019) [57] in China. Whereas, a lower rate of resistance to Ethionamide was reported by Tan Y et al., (39.1%, 2017) [58] and Wu X et al., (2.05%, 2019) [59] in China. The higher rate of resistance to Ethionamide observed in this study may be explained by the fact that Ethionamide is a derivative of isonicotinic acid having a similar structural analogue with isoniazid share common pathways that lead to cross-resistance and resistance to INH seemingly will mediate co-resistance to both isoniazid and ethionamide [60,61]. Notwithstanding this fact, all Ethionamide-resistant isolates were identified as either monoresistant or any-resistant to INH in our study. Meanwhile, there appear recent guidelines recommending against its inclusion within the standard second-line anti-TB treatment regimen, Ethionamide is a second-line pro-drug recommended to be conditionally prescribed for the treatment of MDR-TB. Nevertheless, a higher rate of resistance against Ethionamide observed in this study would mean that the use of ethionamide in the second-line treatment regimens may not have therapeutic benefit in this study area.

Although the primary objective of this study was not to validate rapid molecular testing against the reference standard method, we compared the rate of RR-TB detected using GeneXpert MTB/RIF Assay with BD BACTEC™ MGIT™ 960 SIRE liquid culture system for automated drug susceptibility testing. The sensitivity of 100% of GeneXpert MTB/RIF Assay in detecting rifampicin resistance in our study was in line with previous studies reported by Meawed TE et al., (100%; 2016) [62] in Egypt, Jahan H et al., (98.5%; 2016) [63] in Bangladesh, Pandey P et al., (98.6%; 2017) [64] in Nepal and Mekkaoui L et al., (100%; 2021) [65] in Belgium; but higher than the sensitivity reported in previous studies by Tang T et al., (84.0%; 2017) [66] in China and Feliciano CS et al., (94%; 2019) [67] in Brazil. Whereas, the specificity of 94.6% of GeneXpert MTB/RIF Assay in detecting rifampicin resistance in our study was lower than previous study reported by Meawed TE et al., (100%; 2016) [62] in Egypt, Jahan H et al., (100%; 2016) [63] in Bangladesh, Pandey P et al., (100%; 2017) [64] in Nepal, Mekkaoui L et al., (99.2%; 2021) [65] in Belgium and Feliciano CS et al., (98%; 2019) [67] in Brazil; but higher than the specificity reported by Tang T et al., (87.8%; 2017) [66] in China.

In this study, 52.9% (9/17) of cases showed concordance between GeneXpert MTB/RIF Assay and BD BACTEC™ MGIT™ 960 SIRE liquid culture system for automated phenotypic DST in detecting rifampicin resistance. However, a significant proportion, 47.1% (8/17) of discordance rate was observed. This result was lower than the concordance rate reported by Feliciano CS et al., (90.5%; 2019) [67] in Brazil, but comparable with what was reported in the previous studies by Williamson DA et al., (31%; 2012) [68] in New Zealand and Ngabonziza JCS et al., (47.0%; 2020) [69] in Rwanda. In other words, the rate of detection of rifampicin resistance among GeneXpert MTB/RIF Assay and BD BACTEC™ MGIT™ 960 SIRE liquid culture and DST System was determined at 10.9% (17/156) and 5.8% (9/156) respectively.

However, this study’s findings showed comparable sensitivity, but inconsistent specificity with previous similar studies might be due to the nature and limitations of GeneXpert MTB/RIF Assay as a positive test does not necessarily indicate the presence of viable tubercle bacilli, the presence of mutation conferring rifampicin resistance within as well as outside of the RRDR of the rpoB gene, the presence of silent mutations in the rpoB gene, and low bacterial DNA in the clinical specimens contribute to the increased false rifampicin resistance observed in GeneXpert MTB/RIF Assay in this study. Needless to say, the classical GeneXpert MTB/RIF Assay System is playing a critical role in the screening and detection of *M. tuberculosis* in clinical specimens. However, its efficiency in identifying resistance against rifampicin introduces an unannounced high rate of false positive results leading a significant number of patients to be placed on a second-line anti-TB treatment regimen.

This study has some limitations that need to be noted. Firstly, the study was conducted in new presumptive pulmonary TB cases and this finding could not be generalized to other types of TB disease classification. Secondly, we were not able to include PZA susceptibility testing as we encountered shortages of validated and commercially available reagents. Thirdly, we were unable to follow patients in order to look into treatment outcomes due to logistic reasons.

## Conclusion

The rate of MDR-TB among new pulmonary TB cases remained high at fivefold the national and nearly twofold the global estimated rate. The rate of monoresistance against anti-TB drugs was also high. The absence of resistance against recommended second-line anti-TB drugs was quite encouraging. However, the high rate of resistance against Ethionamide would mean that its inclusion in the regimen may not have therapeutic benefit in this geographic area. Furthermore, the use of GeneXpert MTB/RIF Assay alone for the detection of *M. tuberculosis* might introduce 15.7% false positivity and 47.1% false rifampicin resistance leading patients erroneously assigned in the MDR-TB category and placing on an unnecessary second-line anti-TB-treatment regimen. Enhanced validation and progressive harmonization of rapid molecular diagnostics against reference methods are recommended to improve patient outcomes.

## Data Availability

All relevant data are within the manuscript and its Supporting Information files.

## Supporting information

**S1File**. Sociodemographic, culture, and DST raw data xls.

## Acknowledgment

The authors would like to thank the study participants, specimen collectors, and laboratory technologists at St. Peter TB Specialized Hospital in Addis Ababa, Ethiopia. We are also grateful to acknowledge Department of Microbial, Cellular and Molecular Biology, College of Natural and Computational Sciences, Addis Ababa University, Addis Ababa, Ethiopia, Ethiopian Public Health Institute’s National TB Reference Laboratory, Addis Ababa, Ethiopia, and St. Paul’s Hospital Millennium Medical College in Addis Ababa, Ethiopia.

